# bistro: An R package for vector bloodmeal identification by short tandem repeat overlap

**DOI:** 10.1101/2023.09.14.23295566

**Authors:** Zena Lapp, Lucy Abel, Judith Mangeni, Andrew A. Obala, Wendy O’Meara, Steve M. Taylor, Christine F. Markwalter

## Abstract

1. Measuring vector-human contact in a natural setting can inform precise targeting of interventions to interrupt transmission of vector-borne diseases. One approach is to directly match human DNA in vector bloodmeals to the individuals who were bitten using genotype panels of discriminative short tandem repeats (STRs). Existing methods for matching STR profiles in bloodmeals to the people bitten preclude the ability to match most incomplete profiles and multi-source bloodmeals to bitten individuals.
2. We developed bistro, an R package that implements 3 preexisting STR matching methods as well as the package’s namesake, bistro, a new algorithm described here. bistro employs forensic analysis methods to calculate likelihood ratios and match human STR profiles in bloodmeals to people using a dynamic threshold. We evaluated the algorithm’s accuracy and compared it to existing matching approaches using a publicly-available panel of 188 single-source and 100 multi-source samples containing DNA from 50 known human sources. Then we applied it to match 777 newly field-collected mosquito bloodmeals to a database of 645 people.
3. The R package implements four STR matching algorithms in user-friendly functions with clear documentation. bistro correctly matched 99% (184/185) of profiles in single-source samples, and 63% (225/359) of profiles from multi-source samples, resulting in a sensitivity of 0.75 (vs < 0.51 for other algorithms). The specificity of bistro was 0.9998 (vs. 1 for other algorithms). Furthermore, bistro identified 80% (729/909) of all possible matches for field-derived mosquitoes, yielding 1.4x more matches than existing algorithms.
4. bistro identifies more correct bloodmeal-human matches than existing approaches, enabling more accurate and robust analyses of vector-human contact in natural settings. The bistro R package and corresponding documentation allow for straightforward uptake of this algorithm by others.

## Introduction

Vector-borne diseases cause over 700,000 deaths each year (WHO, 2020). Understanding vector biting behavior in a natural setting can inform precise targeting of interventions to efficiently interrupt transmission. One approach is to identify human factors associated with increased vector biting by matching the human DNA in vector bloodmeals to the individuals who were bitten. Analogous to forensic DNA analysis, this can be done by genotyping panels of short tandem repeats (STRs) in a human population and in vector bloodmeals.

STR genotyping has been used to profile human bloodmeals in sandflies (Inbar et al., 2016), lice (Mumcuoglu et al., 2004), and mosquitoes (Soremekun et al., 2004; Gonçalves et al., 2017; Keven et al., 2021; Mbewe et al., 2023). Existing methods for matching STR profiles in bloodmeals to the people bitten require exact, or near-exact, matching of all alleles at all loci between a bloodmeal and the person bitten. This precludes the ability to match most incomplete locus profiles, and any multi-source bloodmeals with more than two alleles at a locus, to bitten individuals. While there will inevitably be incomplete and multi-source bloodmeals that cannot be confidently matched to any person, for forensic applications, there are robust and well-established methods for matching evidence from incomplete and multi-source samples to persons of interest. These methods provide a likelihood ratio that the DNA of a person of interest is in the evidence sample (vs. a random contributor), which for legal proceedings is typically interpreted with extremely high stringency. Less stringency is needed for matching vector bloodmeals to bitten individuals; in this research context, a small sacrifice in specificity, which would be unacceptable for forensic analysis, can provide valuable gains in sensitivity that lead to useful insights into disease transmission epidemiology.

We adapted forensic analytic approaches to develop bistro (bloodmeal identification by STR overlap), a user-friendly R package that enables matching of bloodmeal STR profiles to people based on log10 likelihood ratios (log10LRs). The underlying algorithm attempts to match not only complete STR profiles from single-source bloodmeals, but also incomplete STR profiles and multi-source bloodmeals. We evaluated the algorithm’s efficacy using samples containing human DNA from known sources, and applied it to mosquito bloodmeals from unknown sources.

### Matching algorithms

While matching people to bloodmeals is not a new concept, to our knowledge no software packages exist that provide user-friendly implementations of bloodmeal-human matching algorithms. Thus, the bistro R package implements not only our newly proposed bistro algorithm, but also three pre-existing methods for matching STR profiles: exact (Soremekun et al., 2004), similarity (Keven et al., 2021; Mbewe et al., 2023), and static threshold (Bleka et al., 2019) (**Table 1**). Exact and similarity matching have been used to match mosquito bloodmeals to people bitten, while static threshold matching is commonly used in forensics. Below, we describe each of the algorithms before explaining their implementation.

**Table 1:** Comparison of matching algorithms.

| Method | Match samples with incomplete profiles | Match samples predicted to be multi-source | Threshold | Use case | Reference |
| --- | --- | --- | --- | --- | --- |
| Exact | No | No | Complete identity | Bloodmeal matching | (Soremekun et al., 2004) |
| Similarity | Limited | No | Highest human similarity | Bloodmeal matching | (Keven et al., 2021; Mbewe et al., 2023) |
| Static threshold | Yes | Yes | Static log10LR | Forensics | (Bleka et al., 2017, 2019) |
| Bistro | Yes | Yes | Dynamic log10LR | Bloodmeal matching | This paper |

### Exact matching

The complete sample STR profile must match the complete human profile.

### Similarity matching

The proportion of exact locus matches between each sample and human is identified, and any similarities above maximum pairwise similarity values of the human database are matches.

### Static threshold matching

Matches are bloodmeal-human pairs with a log10LR above a user-defined threshold.

### bistro matching

To identify human contributors to a bloodmeal, bistro uses pairwise bloodmeal-human log10LRs (**Figure 1**). The log10LRs compare the likelihood that the DNA of a person is in the bloodmeal sample (vs. a random contributor). To be considered a potential match, the likelihood ratio must be above 1.5 (i.e. 30x greater likelihood). For each bloodmeal with at least 1 log10LR ≥1.5, we initialize a set of matches using a minimum log10LR threshold of ⌊*max(log10LR*2)*⌋/*2*. We then decrement the threshold by 0.5 until the number of matches is ≥ the estimated number of contributors (NOC) and the matches identified by the current threshold are identical to the matches identified by the previous threshold. The most confident match set is considered to be the one that was identified by at least two thresholds and where the number of bloodmeal-human pairs is maximized under the constraint that there are ≤NOC pairs. The algorithm returns no bloodmeal-human pairs if no confident match sets are identified by a log10LR threshold = 1, or if all match sets have >NOC pairs.

**Figure 1:**
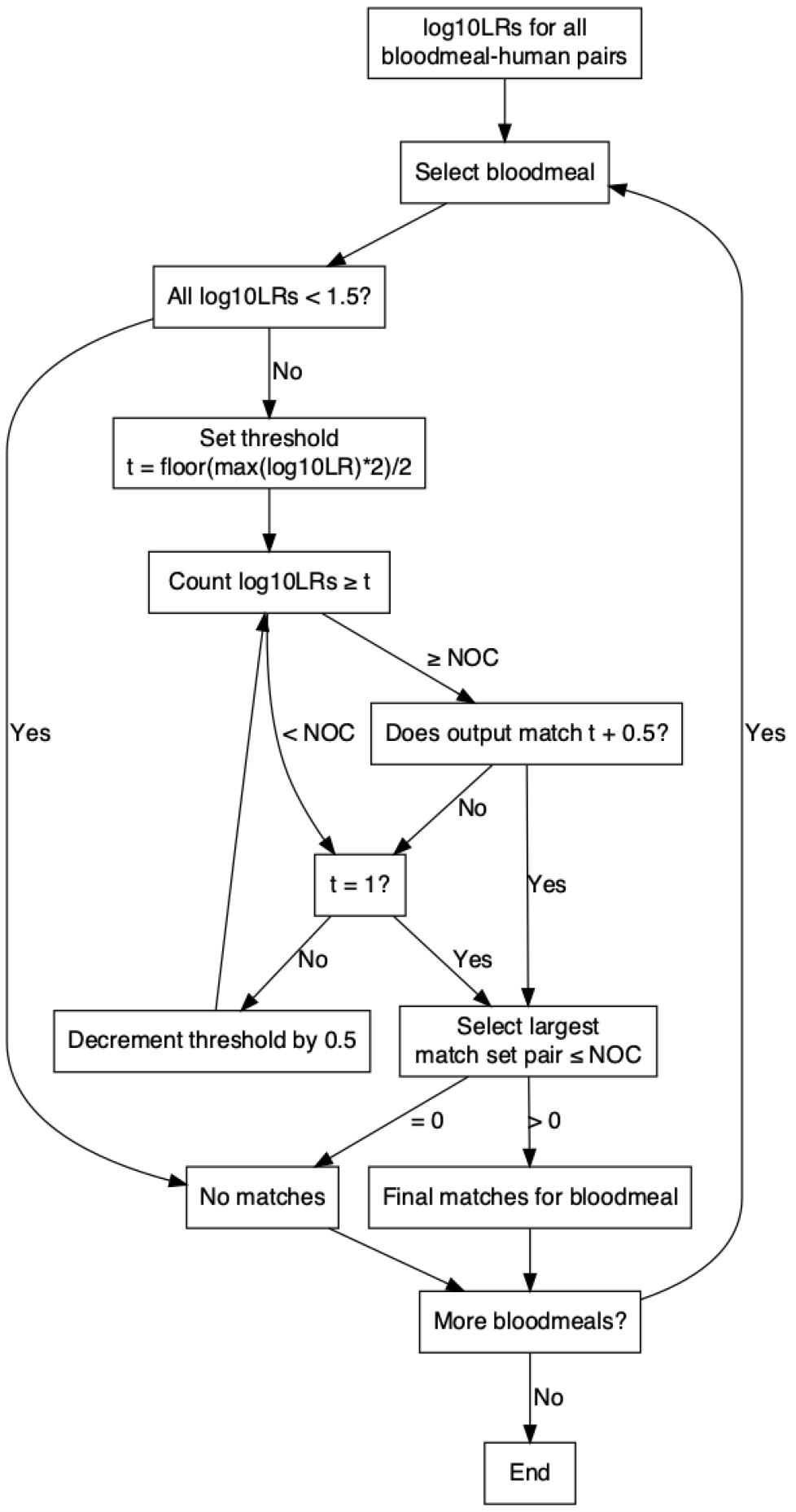
bistro matching algorithm for identifying matches between bloodmeals and people. log10LRs = log10 likelihood ratios; t = threshold; NOC = estimated number of human contributors.

### bistro R package

The open-source bistro R package hosted on GitHub (https://github.com/duke-malaria-collaboratory/bistro), implements the matching algorithms described above and provides examples using a built-in dataset. Documentation on the package website (https://duke-malaria-collaboratory.github.io/bistro/index.html) describes step-by-step instructions for installing the package, running the algorithms, and understanding the output. Below, we briefly describe the implementation of the bistro algorithm.

The bistro function takes as input bloodmeal STR profiles, human STR reference profiles, a bloodmeal STR allele peak detection threshold, the STR genotyping kit name, and optionally human population allele frequencies. Then, it performs the following steps:

1. If no human population allele frequencies are supplied, compute them from the provided human STR profiles.
2. Remove bloodmeal STR alleles below the user-provided peak threshold.
3. Estimate the NOC for each bloodmeal using the equation: ⌈*max(a)/2*⌉, where a is the number of alleles at a locus.
4. Calculate log10LRs for each bloodmeal-human pair using euroformix::contLikSearch() (Oyvind Bleka, 2023) with the parameter defaults in **Table 2**.
5. Use the bistro matching algorithm described above to identify, from the provided human reference database, contributors to each bloodmeal.

**Table 2:** bistro parameters for euroformix::contLikeSearch()

| contLikSearch parameter | Description | bistro default value |
| --- | --- | --- |
| threshT | peak detection threshold in RFU | user-provided |
| NOC | number of contributors | min(NOC, 3) |
| Difftol | tolerance of difference in log likelihood across 2 iterations | 1 |
| modelDegrad | whether to model degradation | TRUE |
| modelBWstutt | whether to model backward stutter | FALSE |
| modelFWstutt | whether to model forward stutter | FALSE |
| prC | allele drop-in probability | 0.05 |
| Lambda | parameter in modeled peak height shifted exponential model | 0.01 |
| nDone | number of optimizations required providing the same log-likelihood | 2 |
| Steptol | minimum allowable relative step length | 0.001 |

The output of the bistro function is a dataset with matches for each bloodmeal, including why there was or was not a match for each.

The current version of bistro (v0.1.1) requires R (≥ 4.0.0) (R Core Team, 2021), depends on euroformix (≥ 4.0.7) (Oyvind Bleka, 2023); and imports codetools (≥ 0.2.19) (Tierney, 2023), R.utils (≥ v2.12.2) (Bengtsson, 2022), and dplyr (≥ 1.1.3), stringr (≥1.5.0), tibble (≥ 3.2.1), and tidyr (≥ 1.3.0) (Wickham et al., 2019).

### Snakemake workflow

We also provide a template Snakemake (Köster & Rahmann, 2012) workflow that enables straightforward parallelization of match identification using bistro (https://github.com/duke-malaria-collaboratory/bistro_pipeline).

## Analysis methods

### Ethics

The human and mosquito data in this study were collected under protocols approved by the IRBs of Duke (Pro00082000) and Moi (IREC\2017\36) Universities.

### Publicly available STR profiles

To evaluate bistro, we used data from the public PROVEDIt database, which provides single- and multi-source, complete and incomplete STR profiles derived from 50 reference individuals (Alfonse et al., 2018). Ratios of sample material from each contributor to multi-source profiles were pre-specified in the dataset. We used profiles originating from untreated samples evaluated with the Identifiler Plus STR kit (28 cycles) on a 3130 Genetic Analyzer with a 10s injection. We evaluated samples with ≥0.1ng template mass and applied a peak height threshold of 200 relative fluorescence units (RFUs), similar to the criteria applied to STR profiles from mosquito bloodmeals below. Population allele frequencies from the revised NIST Population dataset of Autosomal STRs (Hill et al., 2013; Steffen et al., 2017) were used.

### Bloodmeal and human STR profiles from a malaria-endemic region

Indoor resting mosquitoes were collected by aspiration weekly from July 2020 to September 2021 in 75 households across 5 villages in Webuye, Kenya. Bloodfed abdomens (n=1065) were separated from head/thoraces and pressed in filter paper to preserve bloodmeals for STR analysis. Dried blood spot samples (Whatman Protein Saver 903) were collected from each individual who slept in the study households during the study period.

Anopheles bloodmeals and human DBS were extracted using Chelex-100 (Markwalter et al., 2022). Briefly, blood spots were soaked overnight at 4°C in 0.5% saponin before two extractions in 150µl of 6.7% chelex in a 95°C water bath. All extracts were cleaned with Zymo RNA Clean & Concentrator-96 kit using Zymo XL-96 filter plates. Bloodmeal extracts were cleaned and concentrated twice. A template volume of 1µl (human DBS) or 3.5µl (bloodmeal) was added to 10µl amplification reactions using the 10-locus Promega Geneprint10 kit (Promega Cat #B9510). Products (1µl) were analyzed on an Applied Biosystems 3730xl DNA Analyzer with 0.5x injection with a 600 internal lane standard. Alleles were called using Osiris v2.15.1 (NCBI et al., 2021) with a 200 RFU threshold, which eliminated artifacts from no-template control samples.

Population allele frequencies were calculated among people from the cohort (n=645) to more closely represent the allele frequencies of the population present in bloodmeals.

### Data analysis and visualization

We compared the four matching algorithms described above for each dataset. For static threshold matching, we used a threshold of 10. The AMEL marker was not used for methods requiring log10LRs. Data were analyzed and visualized using R v4.2.1 (R Core Team, 2021) and RStudio v2022.12.0+353 (*RStudio*, 2023) with the following packages: tidyverse v2.0.0 (Wickham et al., 2019), ggpubr v0.6.0 (Kassambara, 2018), ggupset v0.3.0 (Ahlmann-Eltze, 2020). Data analysis scripts are on GitHub: https://github.com/duke-malaria-collaboratory/bistro_validation.

## Results

### bistro efficacy

We first evaluated the efficacy of bistro by using it to match people to samples with known sources (**Figure 2A, Table 3**): 188 single-source samples (n=188 sources) and 100 multi-source samples (n=359 sources). For single-source samples, 99% (187/188) of matches were correctly identified, including 3/3 samples with incomplete profiles. For multi-source samples we identified at least one source in each sample and 62% (224/359) of all possible matches, including 5/17 matches from incomplete profiles. Across all samples, those with complete profiles had a higher matching rate (76%) than those with incomplete profiles (40%; **Figure 2B**). Furthermore, within samples, DNA present at higher proportions matched more frequently than DNA present in lower proportions (**Figure 2C**; Wilcoxon p < 0.001).

**Figure 2:**
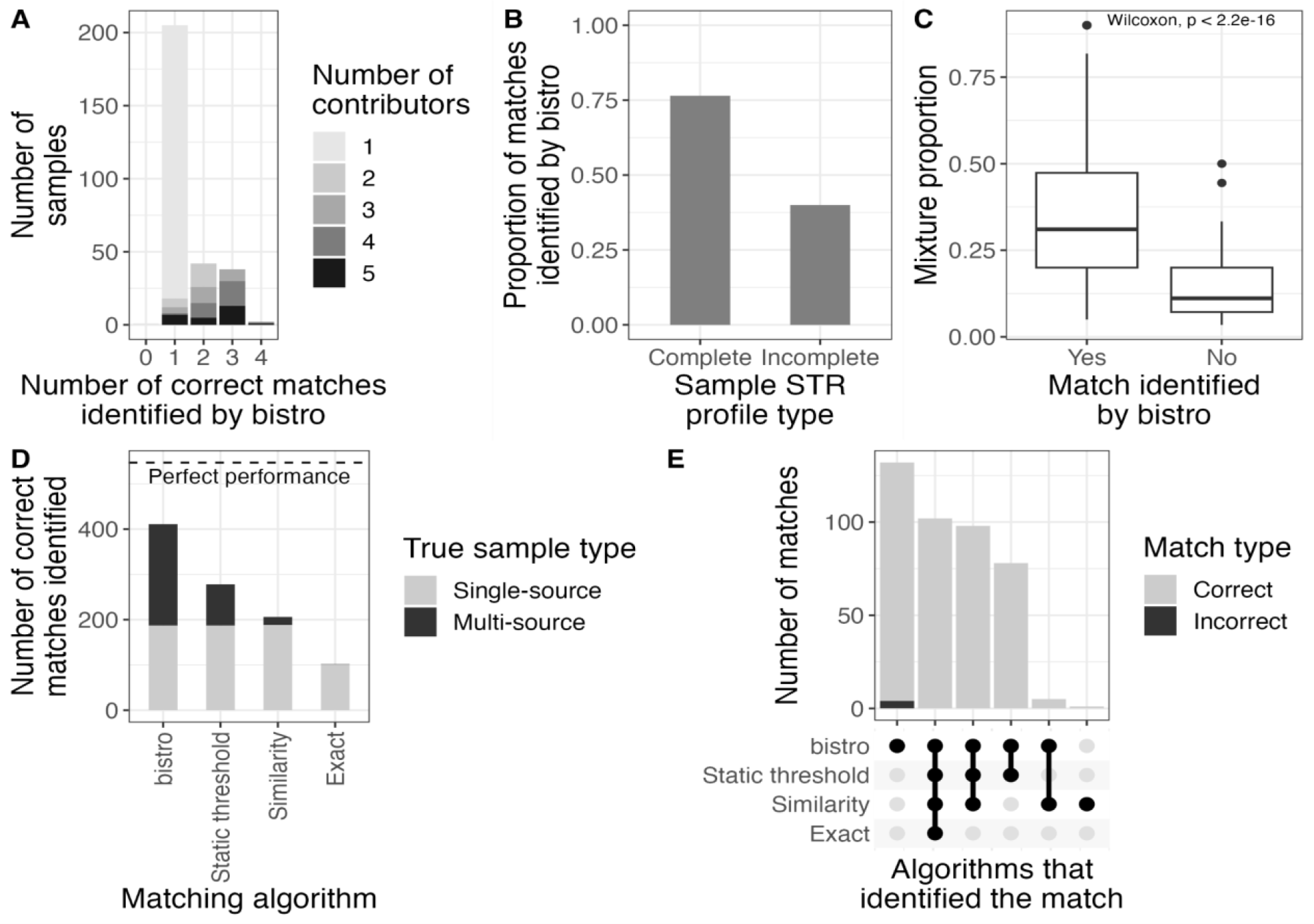
Evaluating bistro using samples with known sources. (A) Number of correct matches for each sample colored by known number of contributors (NOC). (B) Proportion of matches identified separated by whether the STR profile was complete (15 loci present) or not (<15 loci present). The denominator is the total number of possible matches (m_tot_; 547). (C) For multi-source samples, the original DNA mixture proportion of people successfully matched vs. not matched. (D) Total number of matches identified by each algorithm. The dotted line is m_tot_. Color indicates single-vs. multi-source sample based on the known NOC. (E) Number of matches identified by each algorithm.

**Table 3:** Comparison of matching algorithm performance on samples with known sources.

| Algorithm | Single-source | Multi-source |  | All |  |  |
| --- | --- | --- | --- | --- | --- | --- |
| | # correct sources identified | # samples with $\geq 1$ correct source identified | # correct sources identified | # samples with $\geq 1$ correct source identified | # correct sources identified | # incorrect matches |
| Bistro | 187/188 (99%) | 100/100 (100%) | 224/359 (62%) | 287/288 (99.7%) | 411/547 (75%) | 4 |
| Static threshold | 187/188 (99%) | 68/100 (68%) | 91/359 (25%) | 255/288 (89%) | 278/547 (51%) | 0 |
| Similarity | 188/188 (100%) | 18/100 (18%) | 18/359 (5%) | 206/288 (72%) | 206/547 (38%) | 0 |
| Exact | 101/188 (54%) | 1/100 (1%) | 1/359 (0.28%) | 102/288 (35%) | 102/547 (19%) | 0 |

Next, we compared bistro to three other algorithms used for matching: exact, similarity (similarity threshold = 0.375), and static threshold (**Figure 2D-E, Table 3**). bistro returned 4 false positives (specificity = 0.9998), while all other algorithms returned zero false positives (specificity = 1). However, bistro (411 matches, sensitivity=0.75) identified 4x more matches than exact (102 matches, sensitivity=0.19), 2x more matches than similarity (206 matches, sensitivity=0.38), and 1.5x more matches than static threshold matching (278 matches, sensitivity=0.51), yielding much higher sensitivity.

### bistro on mosquito bloodmeals

We next applied bistro to identify matches between a freshly-collected database of human samples (n=645) from Western Kenya and mosquito bloodmeals (n=1065) collected from their households (**Figure 3A, Table 4**). About a quarter (288) of bloodmeals returned no usable peaks from STR typing. Among the 777 bloodmeals that returned peaks, 654 (84%) were estimated to be single-source (n=654 human sources), and 123 (16%) were estimated to be multi-source (n=252 human sources), totalling 906 estimated human sources.

**Figure 3:**
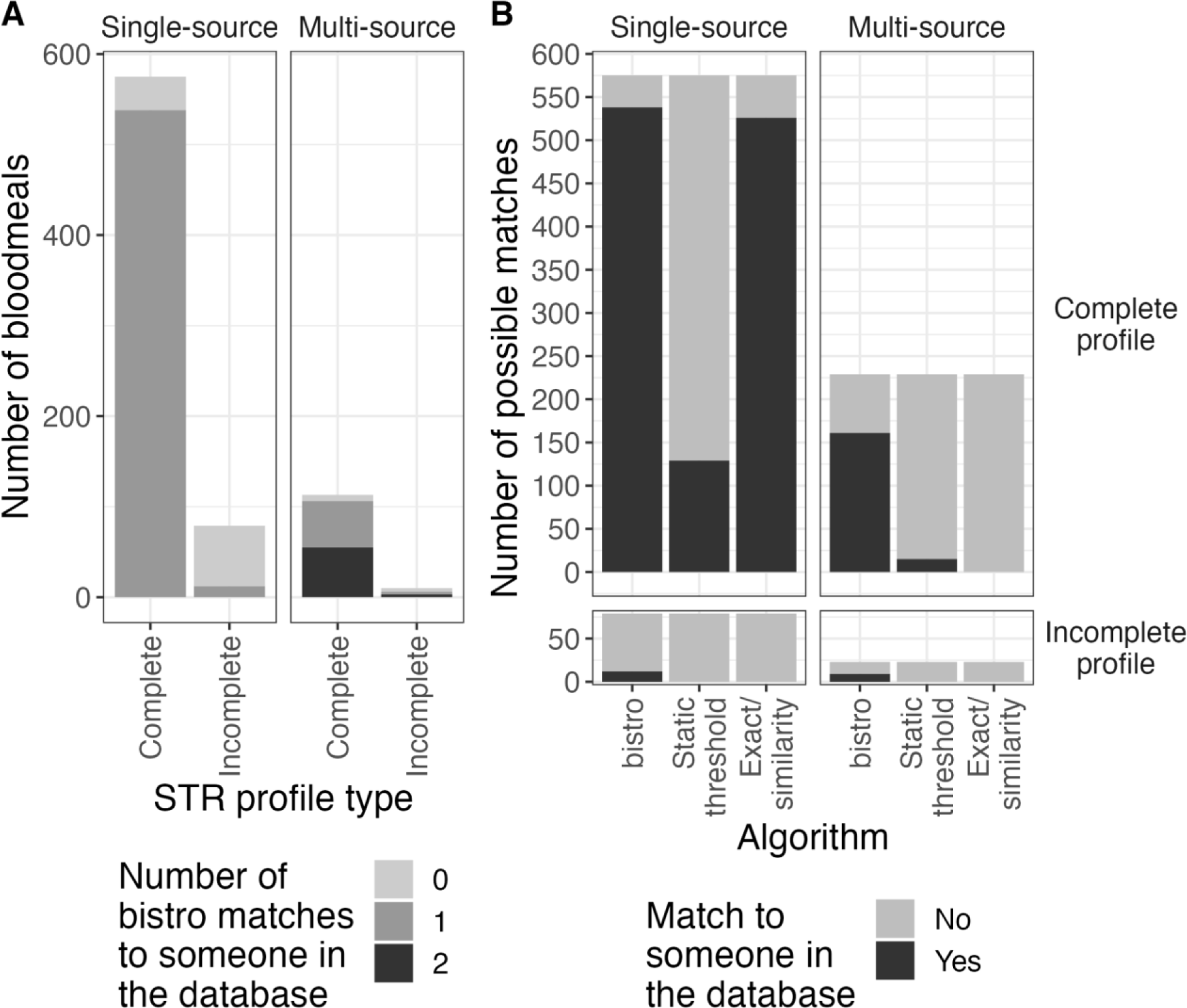
Application of matching algorithms to mosquito bloodmeals. (A) Number of matches identified by bistro for each bloodmeal by single-vs. multi-source bloodmeal (based on estimated NOC) and STR profile completeness. (B) Comparison of matches identified by different matching algorithms by profile completeness and NOC. For this dataset, exact and similarity matching return the same results because the highest similarity value in the cohort was 9/10, requiring an exact match with all 10 STR-typed loci.

**Table 4:** Comparison of matching algorithm performance on Anopheles bloodmeal samples.

| Algorithm | Predicted single-source | Predicted multi-source |  | All |  |
| --- | --- | --- | --- | --- | --- |
| | # sources identified | # samples with $\geq 1$ source identified | # sources identified | # samples with $\geq 1$ source identified | # sources identified |
| bistro | 550/654 (84%) | 112/123 (91%) | 170/252 (67%) | 669/780 (86%) | 720/906 (79%) |
| Static threshold | 129/654 (20%) | 15/123 (12%) | 15/252 (6%) | 530/780 (68%) | 144/906 (16%) |
| Exact/similarity | 526/654 (80%) | 0/123 (0%) | 0/252 (0%) | 526/780 (67%) | 526/906 (58%) |

bistro identified a match to an individual in the database for 94% of complete (644/688) and 20% of incomplete (18/89) bloodmeals. Furthermore, bistro identified two matches for 47% (58/123) of polyallelic profiles thought to represent multi-source bloodmeals. Therefore, we were able to identify 79% (720/906) of all human sources of bloodmeals. In comparison, exact and similarity matching, which are equivalent to each other for these data due to a high maximum similarity value between cohort members (0.9), were only able to identify 58% (526/906) of matches, and static threshold matching was able to identify 16% (144/906) (**Figure 3B, Table 4**). Therefore, bistro identified 1.39x more matches than exact/similarity and 5x more matches than static threshold matching.

## Discussion: strengths and limitations of different matching algorithms

We implemented one novel (bistro) and 3 preexisting (exact, similarity, static threshold) matching algorithms in bistro, an easy-to-use R package. Each of the matching algorithms tested have different capabilities and corresponding strengths and limitations. The benefit of exact matching is that it provides extreme confidence in matches and is not computationally intensive. However, many likely matches are missed with this method because it requires that the estimated NOC equals one and that there are no missing or additional peaks in the bloodmeal STR profile. Similarity matching improves upon this by allowing some non-matching peaks, as long as the proportion of loci that match is greater than the maximum similarity between two STR profiles in a human database. One limitation of this method is that it assumes you have a comprehensive database of who could have been bitten, which may not always be available. Furthermore, as in the bloodmeal data presented here, when cohort members have highly related profiles or when fewer loci are typed, similarity matching becomes identical to exact matching. Static threshold matching, as done in forensic analysis, still returns zero false positives, but nevertheless results in many missing matches that have high log10LRs relative to other samples in the reference database. Additionally, the log10LR magnitude depends on the number of loci typed, so this approach will likely miss more correct matches when evaluating STR panels with fewer loci. To circumvent the limitations of these preexisting algorithms for bloodmeal matching applications, bistro builds upon methods originally developed for forensic analysis to relax the stringency of matches to a degree suitable for research purposes. Since matching bloodmeals in the context of scientific research does not require such high levels of certainty as in forensics, after calculating likelihood ratios for each mosquito-human pair, bistro determines an optimal log10LR threshold for each individual mosquito. This avoids missing true matches with lower log10LRs. While this introduces some false positives, specificity remains high while increasing sensitivity relative to the other algorithms described here. Thus, bistro allows matching of more bloodmeals to people bitten than preexisting methods, providing more matches for downstream analyses related to vector biting and disease transmission. A user-friendly implementation of all four algorithms is provided as an R package with extensive documentation.

## Acknowledgements

This work was supported by the National Institute of Allergy and Infectious Diseases [R01AI146849 to W.P.O. and S.M.T.]. C.F.M. was supported by F32AI149950 and K01AI175527. We appreciate the study implementation skills of project managers and field technicians in Webuye and Eldoret: J. Kipkoech Kirui, I. Khaoya, L. Marango, E. Mukeli, E. Nalianya, J. Namae, L. Nukewa, E. Wamalwa, and A. Wekesa. We thank S. Langdon (DNA Analysis Core, Department of Immunology), B. Freedman, J. Grassia, J. DeCurzio, and T. Thane (each of Duke University) for their help with laboratory samples. Ultimately, we are indebted to the people who participated in our study.

## Data availability

PROVEDIt data are available at https://doi.org/10.1016/j.fsigen.2017.10.006 (Alfonse et al., 2018). Kenyan STR profiles are unshareable.

## Author contributions

CM, ZL, ST, and WPO conceptualized the project. CM and ZL performed methodology and software development, formal analysis, investigation, data curation, original draft writing, and visualization. LA managed the project. JM, AA, WPO, and ST supervised the project and acquired funding. All authors reviewed and edited the manuscript. Our study includes scientists based in the country where the example mosquito samples were collected. All authors contributed intellectually to the research and study design. We cited literature published by scientists from the region.

